# The association between parental, obstetric, socioeconomic, and lifestyle factors and the development of childhood obesity: A Danish Cohort Study

**DOI:** 10.1101/2023.11.23.23298961

**Authors:** Elizabeth la Cour Christiansen, Ida Näslund Thagaard, Paula L. Hedley, Majbrit Johanne Lautrup Hansen, Christine Frithioff-Bøjsøe, Torben Larsen, Jens-Christian Holm, Michael Christiansen, Lone Krebs

**Affiliations:** Dept. Of Gynecology and Obstetrics, Copenhagen University Hospital, Amager and Hvidovre Hospital, Copenhagen, Denmark; Dept. of Obstetrics and Gynecology, Copenhagen University Hospital – Holbæk, Holbæk, Denmark; Dept. of Gynecology and Obstetrics, Copenhagen University Hospital, Nordsjælland Hospital, Hillerød, Denmark; Dept. for Congenital disorders, Statens Serum Institut; Brazen Bio, Los Angeles, California, USA; The Children Obesity Clinic, Accredited European Centre of Obesity Management, Dept. of Pediatrics, Copenhagen University Hospital – Holbæk, Holbæk, Denmark; Novo Nordisk Foundation Center for Basic Metabolic Research, Faculty of Health and Medical Sciences, University of Copenhagen, Copenhagen, Denmark; Faculty of Health and Medical Sciences, University of Copenhagen, Copenhagen, Denmark; Department of Biomedical Sciences, University of Copenhagen, Copenhagen, Denmark; Dept. of Clinical Medicine, University of Copenhagen, Copenhagen, Denmark

**Keywords:** Birthweight, Body Mass Index, Breast Feeding, Childhood Obesity, Maternal Obesity

## Abstract

**Background:** Childhood obesity is a multifactorial disease with complex etiology. Obstetrical factors are seldom taken into considerations.

**Objectives:** To investigate the association between parental, obstetric and lifestyle characteristics, and childhood overweight and obesity.

**Methods:** This retrospective cohort study evaluated associations between birthweight, pre-pregnancy BMI, birth mode, paternal BMI, family history of obesity, parental status, and maternal socioeconomic status and the outcome variable childhood overweight and obesity using logistic regression. Data regarding parental and childhood characteristics were collected through self-administered questionnaires, and obstetric information was retrieved from the Danish Medical Birth Registry.

**Results:** The incidence of childhood overweight and obesity was 11.3 % at a median (IQR) age of 6.51 years (IQR = 2.84). In obese mothers and children who were macrosomic at birth (birthweight ≥4,500 g), the incidence was 21.6% and 23.4%, respectively.

Risk factors for childhood overweight and obesity were macrosomia, (aOR 2.34, 95% CI 1.24-2.19), maternal- and (aOR 2.48, 95% CI 1.78-3.45) paternal overweight and obesity (aOR 2.17, 95% CI 1.44-3.34) and birthweight z-score (aOR 1.13, 95% CI 1.03-1.23). Combining maternal obesity and a macrosomic child gave the highest risk (aOR 7.49, 95% CI 2.05-24.86) Other predictors were divorced-(aOR 2.04, 95% CI 1.13-3.57) and living as a single parents (aOR 3.80, 95% CI 1.31-10.16).

**Conclusions:** Macrosomia combined with maternal obesity was the strongest risk factor for childhood overweight and obesity. Other individual risk factors are parental obesity and socioeconomic factors. This supports the role of lifestyle modification, education-based policies and interventions to prevent high birth weight in counteracting childhood obesity.

## Background

There is a pandemic of obesity, affecting the health of both children and adults. Maternal pre-pregnancy overweight and obesity has been found to influence birthweight, weight later in life, and the risk of the development of adult cardiometabolic disorders.^1^ In Denmark, 22.3% of women of reproductive age (16-44 years old) are overweight (Body mass index (BMI) ≥25 kg/m^2^ to <30 kg/m^2^) and 14.3% are obese (BMI ≥30 kg/m^2^).^2^ Among preschool children, the rate of overweight and obesity is around 10%.^3^

The potential consequences of maternal obesity for pregnancy and birth are multiple.^1^ However, the mechanisms by which maternal BMI influences birth weight, postnatal growth, and the occurrence of metabolic disease later in the child’s life course have not been fully elucidated. It is clear that intrauterine conditions have a major influence on the metabolic development of the child in a life-long perspective. High maternal pre-pregnancy BMI is associated with an elevated risk of the child being large-for-gestational-age (LGA)^4^, having high birth weight (>4,000g)^5^, increased neonatal body fat percentage^6^, overweight and obesity in infancy and childhood^4^, and higher BMI and body fat percentage in adulthood.^1,7^ Furthermore, birth weight may be more predictive of overweight and obesity in infancy than maternal BMI^13^ and several studies have found that high birthweight is independently associated with the development of childhood overweight and obesity.^14^ Childhood obesity is a complex and multifactorial condition; consequently, studies examining this condition routinely adjust for some of the factors known to be involved, such as maternal BMI and birthweight.^15,16^ However, most studies do not adjust for birth mode and paternal BMI.^15,18^

The aim of this study is to investigate the association between several parental, obstetric and lifestyle characteristics and childhood overweight and obesity.

## METHODS

This study is based on a retrospective cohort including 11,704 mothers of 13,513 children born at Holbaek and Slagelse Hospitals in the Region of Zealand of Denmark between January 1, 2006 and December 31, 2011. All mothers who attended their first prenatal routine visit at the ultrasound unit at Holbaek Hospital, around 12 weeks of gestation, were included in the cohort. Clinical data related to pregnancy and birth was retrieved from the Danish Medical Birth Registry and the Astraia Database. The Danish Medical Birth Registry contains information on all births in Denmark including maternal BMI, smoking in pregnancy, parity, gestational age (GA), sex of the child, birth weight, birth mode, and pregnancy and birth complications. The Astraia Database contains information on ultrasound scans, ethnicity, biochemical results, and medical history.

A questionnaire, designed for the study, regarding information on child growth, diseases, eating habits, family history and structure, parental anthropometry, socioeconomic status, and use of medication (Supplementary Material) was sent to 11,704 mothers from the cohort between 2012 and 2016. Mothers who did not respond in the first round were contacted a second time, 2,339 (20.0%) completed questionnaires were returned (Figure 1). Data from the questionnaires were entered into an SPSS database by two investigators and merged with data from the Astraia Database and the Danish Medical Birth Registry. Discrepancies were addressed by referring back to the questionnaires in order to correct data which had been incorrectly inputted or formatted. We subsequently excluded multifetal pregnancies, preterm births, stillborn or neonatal deaths, children with invalid civil registration numbers, mismatched mother child pairs, and mothers with no information on pre-pregnancy BMI. Furthermore, children with missing data on GA, height and weight measures or duration of breastfeeding were excluded (Figure 1). Analyses were performed on data from 1,448 children and mothers (Figure 1), younger siblings were excluded.

**Figure 1.**
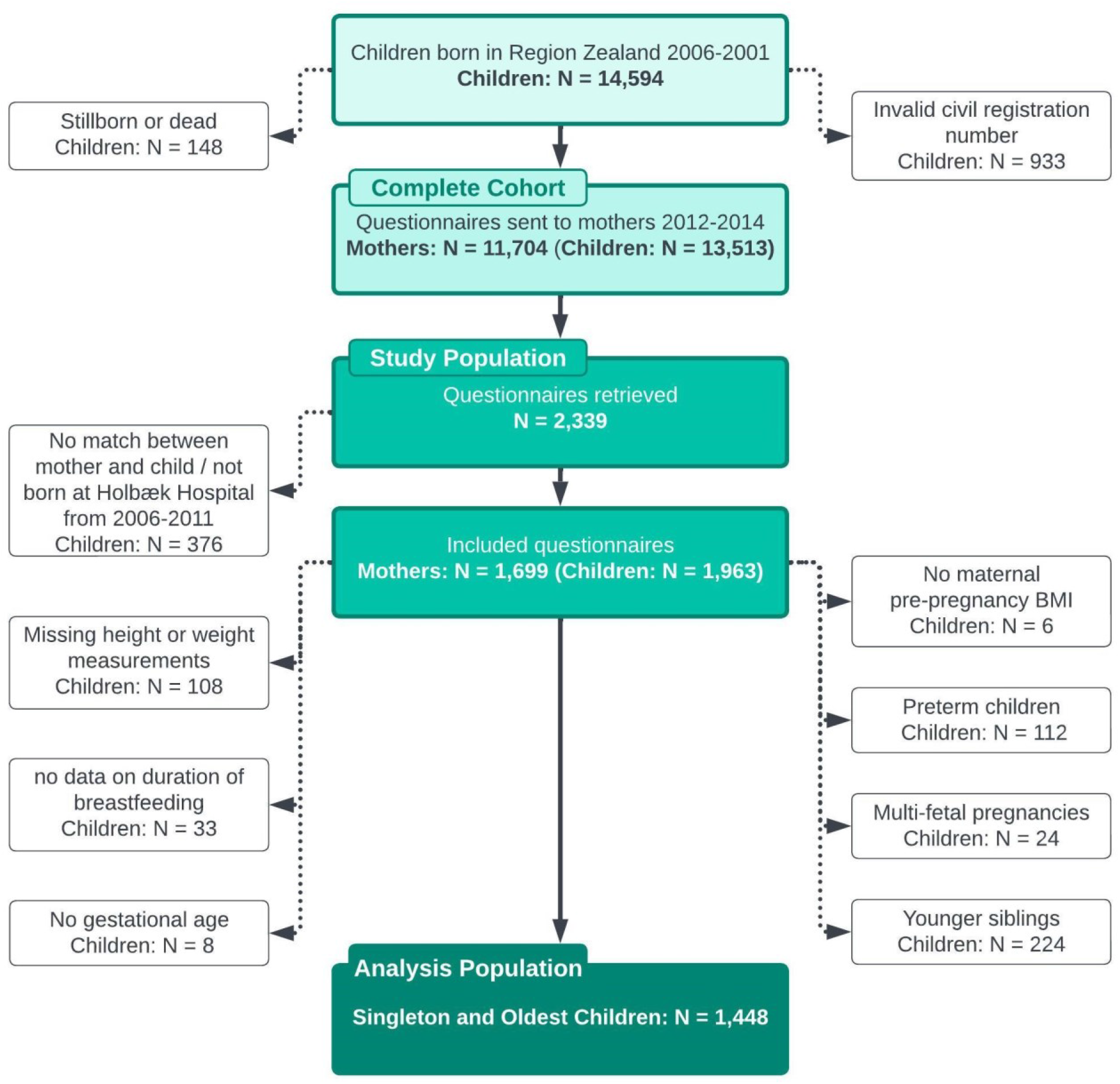
Flow chart describing the inclusion of mother-child pairs in the retrospective cohort.

Adult BMI was calculated as weight in kilograms divided by height in meters squared (kg/m^2^). BMI categorized into groups, according to WHO standards; Underweight (BMI <18,5 kg/m^2^), normal (BMI ≥18,5 kg/m^2^ to <25 kg/m^2^), overweight (BMI ≥25 kg/m^2^ to <30 kg/m^2^), moderate (BMI ≥30 kg/m^2^ to <35 kg/m^2^) and severe obese (BMI ≥35 kg/m^2^).^19^ Birth weight was measured in grams and small for gestational age (SGA) was defined according to the Scandinavian growth charts modified from Marsal *et al* defined as a birth weight deviation of ≤-22% and appropriate for gestational age (AGA) as birth weight deviation between >-22% and <22%, and large for gestational age (LGA) as birth weight deviation ≥22%.^20^ Low birth weight (LBW) was defined as birth weight < 3,000 g, and Macrosomia was defined as birth weight > 4,500 g.

Age and sex specific BMI z-scores for the children were calculated based on height and weight measurements completed by the general practitioner at the preventive health checks at five weeks, five months, one year, two years, three years, four years, and five years of age. Mothers gave information about the exact dates of the health checks and the corresponding weight and height of their child. However, for 264 of the children the dates were missing, we therefore estimated, based on their birth dates, the most likely dates for the planned preventive health check at five weeks, five months, etc. The sex and age specific BMI z-score for each of the measurements was calculated by converting BMI of the child (kg/m^2^) into a normal distribution using the LMS-method comparing the data with the Danish Reference population.^21,22^ Overweight and obesity was defined as a BMI > 90^th^ percentile corresponding to a BMI z-score of ≥1.28. In this study, we used the BMI z-score for the last recorded measurements for each child.

The following covariates regarding pregnancy were considered and categorized as dichotomous variables; smoking, medical diseases, allergies, use of antibiotics or other medications, family history of overweight and obesity (mother, father, siblings, and/or grandparents), and complications in pregnancy. Complications in pregnancy included gestational diabetes (GDM), and hypertensive disorders. Birth mode was categorized as vaginal delivery, cesarean section before labor or cesarean section during labor. Parental socio-economic status was categorized based on current occupation into three groups of employment and one group of unemployment. The employed groups were further categorized based on education; higher, lower, and unclassified. Students and mothers on maternity leave were categorized into the category they headed for or their usual occupation respectively. 93 parents could not be categorized due to unknown current occupation. Duration of breastfeeding was a continuous variable measured in months. Current eating patterns were reported regarding how often the child was eating “fast food” and drinking soda with or without sugar. Current living situation of the parents were categorized as married, cohabiting, divorced and single parents. Maternal ancestry was dichotomized into women of European or non-European descent. The study was reported according to the STROBE guideline.

## STATISTICAL ANALYSES

The characteristics of the study population were described by the count, percentage, mean, standard deviation (SD), median, and interquartile range (IQR). Normality was examined for each variable using visual inspection of residual plots and the Shapiro-Wilk W test for normality. Comparisons of characteristics between normal weight children and children with overweight and obesity were done using Pearson’s chi-squared test or Fisher’s exact test for categorical variables and Students T-test or Mann-Whitney U test for continuous variables.

Multivariable logistic regression was used to assess the association between predictor variables and overweight and obesity in childhood while adjusting for the effect of all included variables, returning adjusted odds ratios (aOR) with 95 % confidence intervals (CI). Variables were excluded from the multivariate logistic regression model if they were demonstrated to be correlated with other variables in the model, in which case the correlated variable with highest complexity were included.

The statistical analyzes were performed using R version 4.2.3.

## ETHICAL APPROVAL

The study was approved by the Regional Scientific Ethics Committee of the Region of Zealand (No: SJ-335) date of approval: 3^rd^ April 2013. Data handling and storage was approved by the Danish Data Protection Agency (No: SJ-HO-01), date of approval: 6^th^ July 2013.

## RESULTS

The overall incidence of childhood overweight and obesity was 11.3% at a median age of 6.51 years (IQR = 2.84). Among mothers with pre-pregnancy BMI ≥ 30 kg/m^2^ the incidence of childhood overweight and obesity among their offspring was 21.6%. In children born macrosomic the incidence was 23.4%. 55.5% of the children with overweight and obesity had a mother with a pre-pregnancy BMI ≥25kg/m^2^, whereas 33.5% of the children with normal weight had a mother with a pre-pregnancy BMI ≥25kg/m^2^.

Univariate analyses showed that childhood overweight and obesity was related to maternal and paternal BMI, a lower education level, a family history of obesity and parents divorced and not living together, maternal smoking at time of questionnaire, high birthweight, median duration of total breastfeeding (months), and consumption of beverages with artificial sweeteners in childhood (Table 1).

**Table 1.** Characteristics of children in different end-weight z-score groups.

|  | All<br>children<br>n = 1,448 | Normal<br>weight<br>children<br>n = 1,284<br>Z-score <<br>1.28 | Overweight<br>and obese<br>children<br>n = 164<br>Z-score<br>≥1.28 |
| --- | --- | --- | --- |
| Child age (years) - final measurement, median (IQR) | 6.20 (2.61) | 6.18 (2.64) | 6.51 (2.84) |
| Child Z-score - final measurement, median (IQR) | -0.04<br>(1.44) | -0.19 (1.27) | 1.70 (0.56) |
| Pregnancy Characteristics |  |  |  |
| Mothers of European ancestry, n (%) | 1,307<br>(90.3) | 1,157 (90.1) | 150 (91.5) |
| Pre-pregnancy BMI, median (IQR) | 23.44<br>(5.70) | 23.17 (5.32) | 25.48 (7.77) |
| Pre-pregnancy BMI ≥25kg/m<sup>2</sup> [Yes], n (%) | 521 (36.0) | 430 (33.5) | 91 (55.5) |
| Mother's age at delivery, mean (sd) | 31.32<br>(4.69) | 31.37(4.68) | 30.89 (4.70) |
| Parity, n(%) |  |  |  |
| 1 | 687 (47.4) | 619 (48.2) | 68 (41.5) |
| 2 | 497 (34.3) | 433 (33.7) | 64 (39.0) |
| >2 | 264 (18.2) | 232 (18.1) | 32 (19.5) |
| Smoking cigarettes during pregnancy, n (%) | 133 (9.2) | 113 (8.8) | 20 (12.2) |
| Hypertensive disorder during pregnancy, n (%) | 34 (2.3) | 33 (2.6) | 1 (0.6) |
| Diabetes during pregnancy, n(%) | 33 (2.3) | 30 (2.3) | 3 (1.8) |
| Gestational age (days) at birth, median (IQR) | 281 (13) | 281 (13) | 281 (15) |
| Birth mode, n (%) |  |  |  |
| vaginal delivery | 1,130<br>(78.0) | 994 (77.4) | 136 (82.9) |
| caesarean section, before labour | 159 (11.0) | 141 (11.0) | 18 (11.0) |
| caesarean section, during labour | 159 (11.0) | 149 (11.6) | 10 (6.1) |
| Sex, n (%) |  |  |  |
| female | 717 (49.5) | 630 (49.1) | 87 (53.0) |
| male | 731 (50.5) | 654 (50.9) | 77 (47.0) |
| Birthweight Z-score, median (IQR) | -0.01<br>(2.75) | -0.08 (2.74) | 0.32 (2.70) |
| Birthweight categories, n (%) |  |  |  |
| LBW (< 3,000g) | 165 (11.4) | 157 (12.2) | 8 (4.9) |
| ABW (≥ 3,000g, < 4,500g) | 1,219<br>(84.2) | 1,078 (84.0) | 141 (86.0) |
| macrosomia (≥4,500g) | 64 (4.4) | 49 (3.8) | 15 (9.1) |
| Birthweight relative to gestational age groups, n (%) |  |  |  |
| SGA | 48 (3.3) | 47 (3.7) | 1 (0.6) |
| AGA | 1,323<br>(91.4) | 1,176 (91.6) | 147 (89.84) |
| LGA | 77 (5.3) | 61 (4.8) | 16 (9.8) |

| Early Childhood Characteristics |  |  |  |
| --- | --- | --- | --- |
| Breastfed infants, n (%) | 1,341 (92.6) | 1,193 (92.9) | 148 (90.2) |
| Exclusively breastfed (months), median (IQR) | 5 (3) | 5 (3) | 4 (5) |
| Diseases in childhood, n (%) | 223 (15.4) | 190 (14.8) | 33 (20.1) |
| Allergies in childhood, n (%) | 132 (9.3) | 114 (9.0) | 18 (11.1) |
| Use of medicine in childhood, n (%) | 150 (10.4) | 127 (9.9) | 23 (14.0) |
| Use of antibiotics in childhood, n (%) | 1,156 (81.2) | 1,019 (80.7) | 137 (84.6) |
| Fast food habits, n (%) |  |  |  |
| Never | 160 (11.1) | 138 (10.8) | 22 (13.5) |
| 1-2 times per week | 1,080 (74.9) | 962 (75.2) | 118 (72.4) |
| 3-6 times per week | 193 (13.4) | 170 (13.3) | 23 (14.1) |
| 7-10 times per week | 7 (0.5) | 7 (0.5) | 0 (0) |
| > 10 times per week | 2 (0.1) | 2 (0.2) | 0 (0) |
| Consumption of sugary drinks, n (%) |  |  |  |
| Never | 380 (26.4) | 332 (26.0) | 48 (29.6) |
| 1-2 times per week | 810 (56.2) | 725 (56.7) | 85 (52.5) |
| 3-6 times per week | 194 (13.5) | 172 (13.4) | 22 (13.6) |
| Everyday | 57 (0.4) | 50 (3.9) | 7 (4.3) |
| Consumption of sugar-free drinks, n (%) |  |  |  |
| Never | 798 (55.3) | 724 (56.7) | 74 (45.1) |
| 1-2 times per week | 384 (26.6) | 334 (26.1) | 50 (30.5) |
| 3-6 times per week | 176 (12.2) | 150 (11.7) | 26 (15.9) |
| Everyday | 84 (5.8) | 70 (5.5) | 14 (8.5) |
| Parental Characteristics |  |  |  |
| Maternal SES, n (%) |  |  |  |
| Employed, higher education | 803 (56.2) | 724 (57.1) | 79 (48.8) |
| Employed, lower education | 447 (33.4) | 412 (32.5) | 65 (40.1) |
| Employed, unclassified | 96 (6.7) | 83 (6.5) | 13 (8.0) |
| unemployed | 54 (3.8) | 49 (3.9) | 5 (3.1) |
| Paternal SES, n (%) |  |  |  |
| Employed, higher education | 537 (38.7) | 487 (39.5) | 50 (32.7) |
| Employed, lower education | 663 (47.8) | 583 (47.3) | 80 (52.3) |
| Employed, unclassified | 162 (47.8) | 141 (11.4) | 21 (13.7) |
| unemployed | 24 (1.7) | 22 (1.8) | 2 (1.3) |
| Maternal BMI at time of questionnaire, median (IQR) | 23.94 (5.84) | 23.78 (5.50) | 26.50 (7.59) |
| Maternal overweight/obese at time of Questionare, n (%) | 578 (40.2) | 480 (37.7) | 98 (60.1) |
| Maternal BMI change since pregnancy, median (IQR) | 0.37 (2.30) | 0.37 (2.21) | 0.70 (2.97) |
| Maternal BMI change since pregnancy, n (%) |  |  |  |
| no change | 717 (49.9) | 655 (51.4) | 62 (38.0) |
| weight gain (upper quartile) | 360 (25.1) | 308 (24.2) | 52 (31.9) |
| weight loss (lower quartile) | 360 (25.1) | 311 (24.4) | 49 (30.1) |
| Paternal BMI at time of questionnaire, median (IQR) | 25.74<br>(4.48) | 25.54 (4.25) | 27.14 (5.56) |
| Paternal overweight/obese at time of Questionare, n (%) | 799 (59.2) | 684 (57.1) | 115 (75.7) |
| Family members with overweight/obese, n (%) | 585 (41.2) | 492 (39.1) | 93 (57.4) |
| Maternal smoking at time of questionnaire, n (%) | 117 (8.1) | 95 (7.4) | 22 (13.6) |
| Father smoking at time of questionnaire, n (%) | 243 (17) | 208 (16.4) | 35 (21.5) |
| Marital status, n (%) |  |  |  |
| Co-habiting | 260 (18.0) | 229 (17.8) | 31 (19.0) |
| married | 984 (68.0) | 888 (69.2) | 96 (58.9) |
| divorced | 153 (10.6) | 124 (9.7) | 29 (17.8) |
| single parent | 49 (3.4) | 42 (3.3) | 7 (4.3) |

Abbreviations: SD = Standard deviation. IQR = Interquartile range. SGA = Small for gestational age. AGA = Appropriate for gestational age. LGA = Large for gestational age.

Following the removal of correlated predictor variables multivariable logistic regression was used to assess the association with childhood overweight and obesity while adjusting for all other variables in the model (Table 2). Univariable logistic regression of all predictor variables used in the multivariable model showed that developing childhood overweight and obesity was positively associated with pre-pregnancy BMI ≥ 25kg/m^2^, birthweight z-score, regular consumption of sugar-free drinks, employed mothers, with lower educational attainment, the BMI of fathers and family members generally, and parents not living together (divorced or single parent households) (Table 2). Having a cesarean section during labor were negatively associated with the development of childhood overweight and obesity (Table 2). Multivariable logistic regression determined that pre-pregnancy BMI≥25kg/m^2^, the BMI of fathers, birthweight z-score, as well as parents divorced or single parents remained positively associated with childhood overweight and obesity after adjustment for all included variables. Furthermore, having a cesarean section during labor remained negatively associated (Table 2).

**Table 2.**
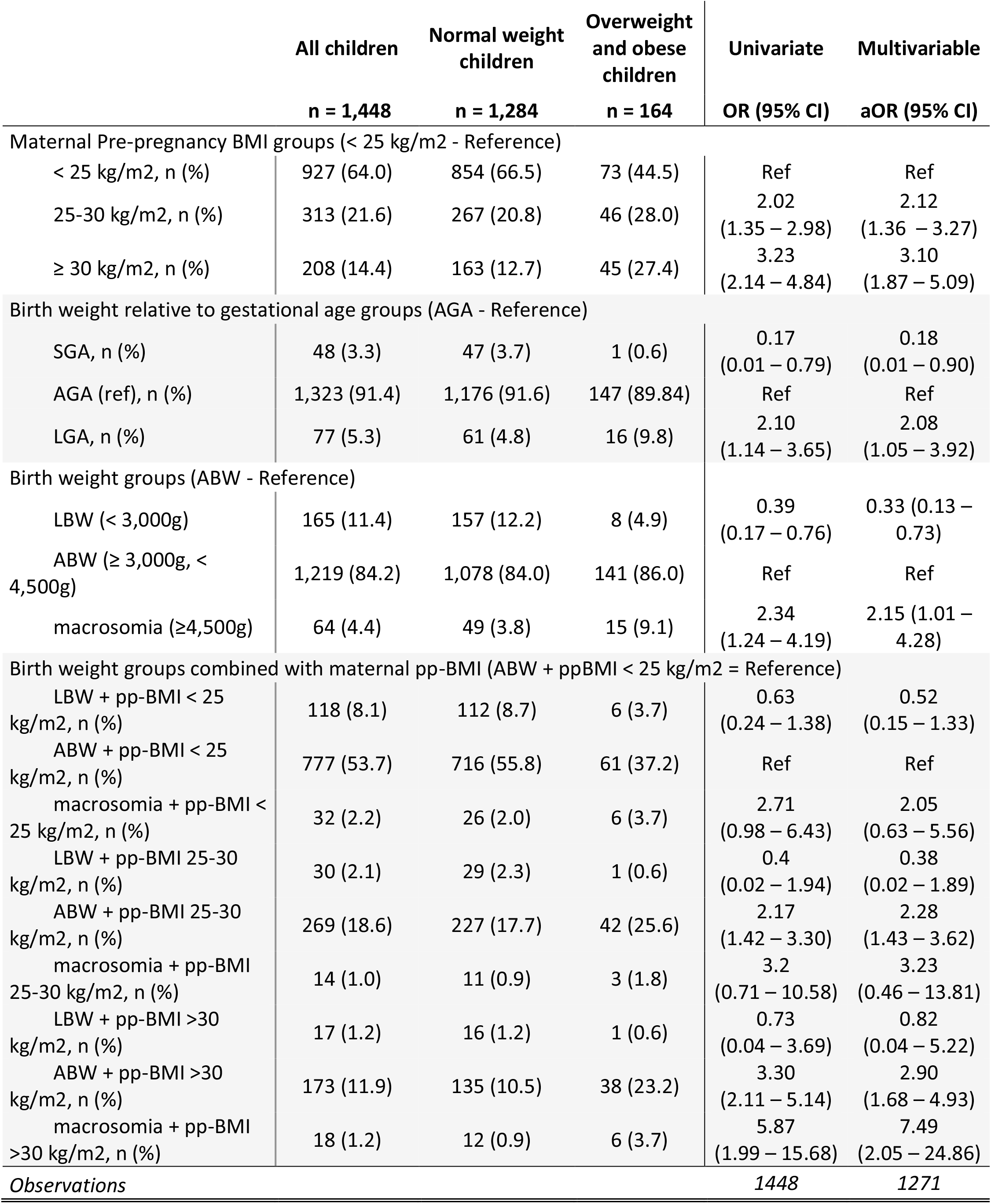
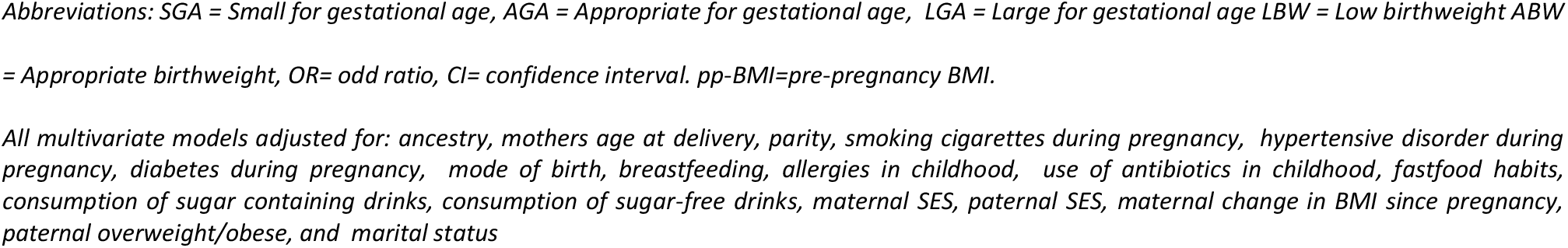
Relative contribution of birth weight and maternal pre-pregnancy BMI to the risk of childhood overweight and obesity.

The relative contribution of birth weight and maternal pre-pregnancy BMI on childhood overweight and obesity is presented in Table 2.

Other significant variables, contributing to childhood overweight and obesity, were a family history of overweight and obesity (aOR 1.63, 95% CI: 1.17-2.48), divorced/single parents (aOR 1.81, 95% CI: 1.20-2.72), lower educational level of the mother (aOR 1.52, 95% CI: 1.07-2.15), and paternal overweight (aOR 1.61, 95% CI: 1.06-2.44) and obesity (aOR 2.60, 95% CI: 1.58-4.24). Cesarean section was protective for childhood overweight and obesity (aOR 0.59, 95% CI: 0.38-0.92) while duration of breast feeding was not (aOR 0.98, 95% CI: 0.95-1.01) (Table 3).

**Table 3.**
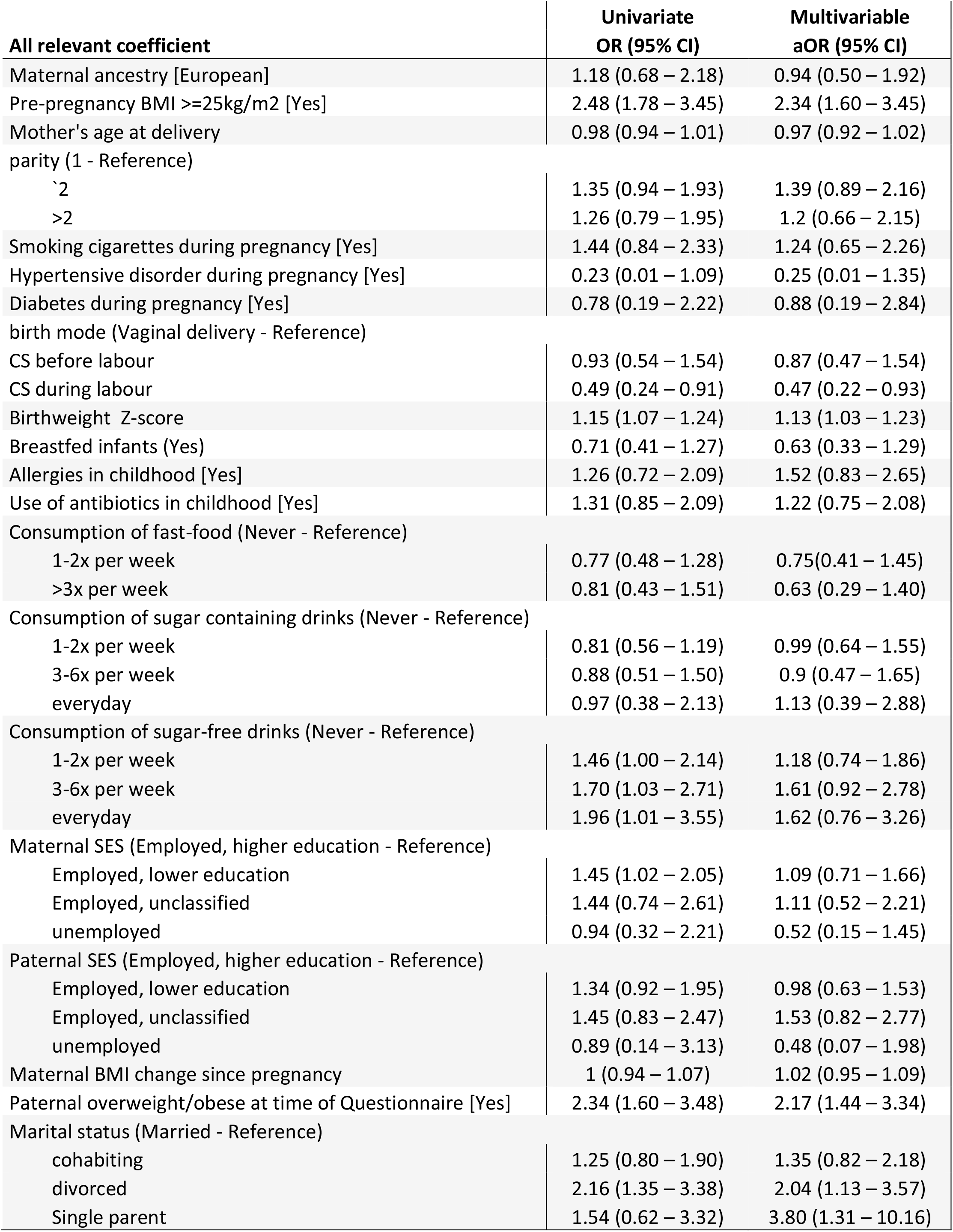
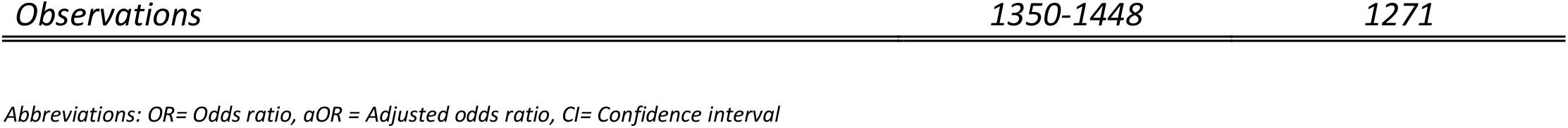
Other factors contributing to childhood overweight and obesity.

Compared to the complete cohort, the responders of the questionnaires had a lower pre-pregnancy BMI, were older, more often non-smokers and of Northern European ancestry, had a lower parity, and delivered infants with higher birth weight when compared to non-responders (Table 4).

**Table 4.**
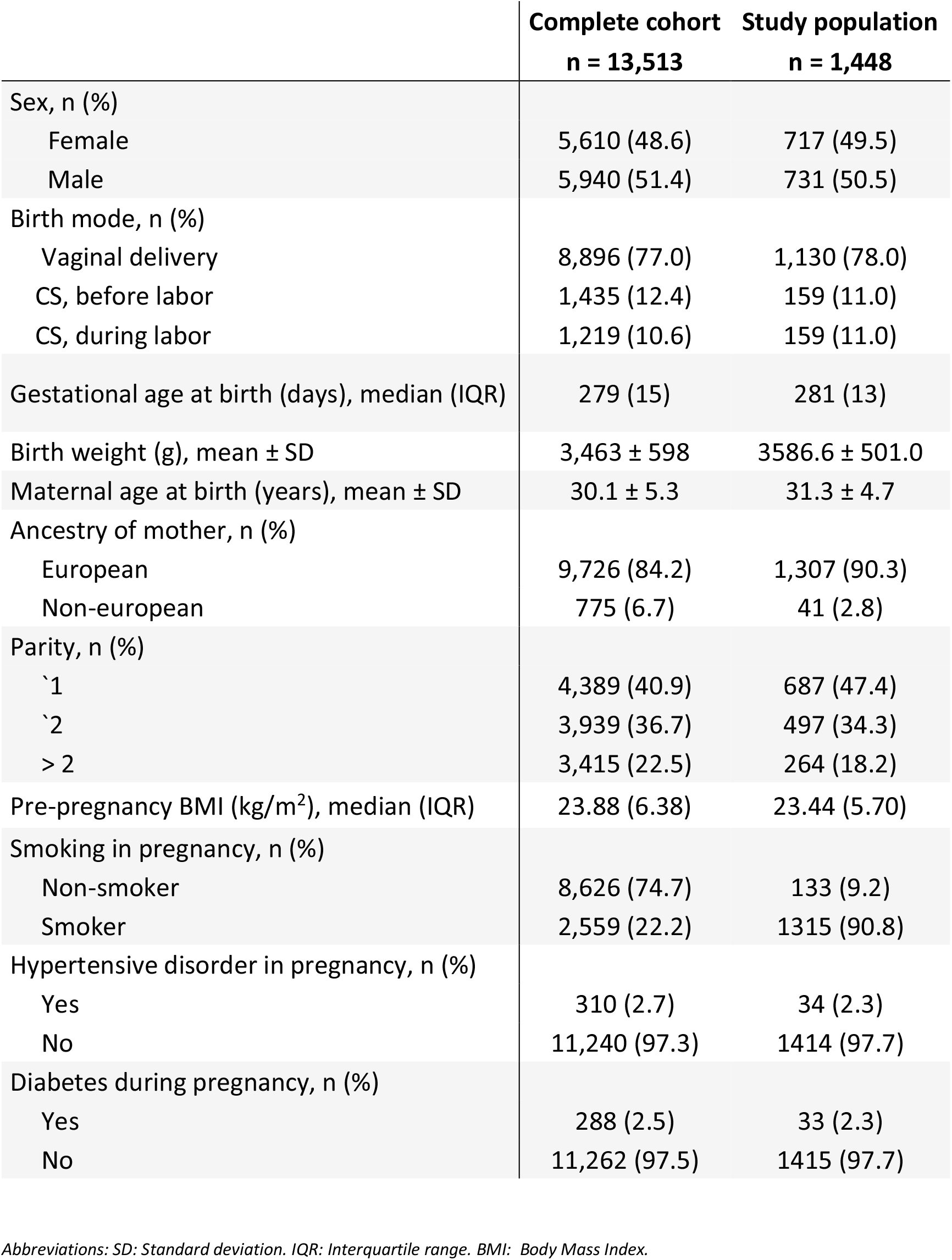
Comparison of the complete cohort and study population.

## Comment

In this study, we found that a combination of maternal obesity and children born macrosmoic was the most pronounced risk factor for childhood overweight and obesity. Maternal obesity was the strongest individual risk factor. After adjustment for confounders, other risk factors were being born macrosomic, paternal overweight and obesity, and parents divorced or single parent. Breast feeding or length of breast feeding had no significant impact.

The association between birthweight and childhood overweight and obesity is well-documented.^14^ However, few studies have evaluated the isolated effect of macrosomia. In accordance with our results, a Chinese study found an OR of 2.7 for developing overweight and obesity in children with macrosomia (defined as birth weight ≥4,000 g), but the follow-up time was only 3 years.^23^ Another Chinese study found a risk of 5.6% for obesity in adulthood when having a birth weight >4,500 g as opposed to a risk of 1.9% if birth weight was 3,500-3,999 g.^24^ A recent Chinese study found a significant, maternal age independent, association between maternal pre-pregnancy BMI and birth of a macrosomic child.^25^

In our study, maternal obesity contributed less to childhood overweight and obesity than macrosomia with a follow up time up to 9.3 years of age. We also found that normal weighted women having a child with AGA had an independent protective effect on developing childhood overweight and obesity. This was also documented by Sauder *et al*, however their follow up period was only five months.^13^

In this study, maternal pre-pregnancy overweight or obesity and high birthweight were independently associated with childhood overweight and obesity. This has been reported previously, but these reports focused on specific ages 3 and 4 years.^26, 27^ They also used different BMI-categories for mothers, and different recordings of maternal BMI as some are measured before conception others at delivery. A study by Gaillard *et al*. showed that maternal obesity enhanced the risk of childhood obesity with an OR of 5.02; our study found a lower contribution (aOR 2.37). The Gaillard study did not adjust for birthweight, which might explain the difference.^8^ Surprisingly, our study did not find an association between SGA and later development of childhood overweight and obesity as suggested by Barker *et al.*^28^ This may be explained by the relatively low prevalence of SGA in our study. The Barker study is based on a cohort from the beginning of the 1900s where the pregnancy exposure profile was very different with regard to nutrition status, physical activity and medical practice; exposure effects on birth weight has been shown to vary with time from the 30’s to 1989.^28^

We found no association with length of breast feeding and overweight and obesity in childhood. Previously, a large systematic review and meta-analysis including 332,297 preschool children found that breast feeding reduced childhood obesity with an OR of 0.83 compared to children who were never breastfed.^15^ The effect depended on the duration of breast feeding i.e., an OR of 0.67 was reported when the duration of breast feeding was > 6 months. In the present study, we found a small trend towards a protective effect of breast feeding, but our sample is probably too small to document a possible positive effect. The prevalence of never breastfed children is low in our study (7.3%), which makes it difficult to evaluate a potential negative effect. Breast feeding is generally popular in Nordic populations, as shown in a Norwegian cohort compared to a British cohort where 90.1% versus 70.0% were breastfed.^30,31^

Reports on the effect of ceasarean sectio on obesity in childhood are contradictory ^32^. We find a protective effect when the ceasarean is performed as an emergency ceasarean during labor. Here the women are in an active state of labor were there are several factors interfering. Other studies have found that elective ceasarean is associated with the elevated risk of childhood obesity not the emergency ceasarean^33^. However, there are few emergency ceasarean in our study precluding robust conclusions.

Other studies have examined combinations of risk factors, since overweight and obesity in childhood is known to be of a complex etiology. Robinson *et al*.^34^ focused on five early-life risk factors, maternal obesity before pregnancy, excessive gestational weight gain, smoking in pregnancy, low vitamin D status in pregnancy, and no or short duration of breastfeeding, and found that the greater the number of risk factors, the higher the risk of childhood overweight and obesity. We were not able to analyze gestational weight gain or vitamin D status in our study. Furthermore, with smoking, as with breastfeeding, we saw a trend, albeit non-significant, towards increased risk of childhood overweight and obesity. A large systematic review and meta-analyses with 84,563 children has found an OR of 1.5 for childhood overweight and obesity when the mother smoked during pregnancy.^35^ Use of antibiotics in infancy has been associated with childhood overweight and obesity, specifically for boys.^36^ The lack of an association in our study may be due to recall bias, as we did not have access to prescription records.

Pregnancy complications such as GDM were not associated with childhood overweight and obesity in our study. Nehring *et al* found GDM to be a risk factor for childhood overweight and obesity^37^ while Gomes *et al*, did not.^38^ The later pointed out the association between late pregnancy dysglycemia, and LGA and childhood overweight and obesity.

When analyzing social factors affecting childhood overweight and obesity, we found that current living situation of the parents affected the risk significantly. This is in accordance with a reported finding of an increased childhood overweight and obesity when parents are divorced.^39^ Parental living situation and socioeconomic factors may be confounders of childhood overweight and obesity, describing the general environment children grow up in. However, complex non-linear interactions of childhood overweight and obesity, such as sociodemographic factors, lifestyle choices, as well as nutrition and exercise, are difficult to exclude.

Nevertheless, a study comparing siblings born prior to and after maternal weight loss, due to bariatric surgery, found that modification of maternal overweight decreased the risk of childhood obesity (age 2-18 years) by 52.0%.^40^ This indicates that maternal pre-pregnancy overweight is a modifiable health factor for the development of childhood overweight and obesity.

The strength of this study is the use of a large and well-described cohort, which contributes to the validity and generalizability of the study. Furthermore, we used international classifications of BMI-categories (WHO-criteria) and end-weight z-score were calculated based on age- and sex specific data from Denmark, with a follow up time up to 9 years of age.

A limitation of the present study is that the low response rate of 20.0% of questionnaires could lead to selection bias. By comparing responders to the complete cohort, we found that they differed by BMI as responders had a lower BMI and were less prone to smoke in pregnancy. Furthermore, the responders had children with a higher birthweight, were more often nulliparous, had higher maternal age at birth, and a higher percentage were of Northern European ancestry. This indicates that the responders had a higher social status. Unfortunately, information regarding social status was not available for the complete cohort. Another limitation is the lack of information on gestational weight gain, since several other studies have shown an increased risk of high birth weight and overweight and obesity in childhood for offspring born to mothers with an excessive gestational weight gain.^4^

This cohort study cannot demonstrate causality, but we have provided evidence for association between childhood obesity and putative causative factors. However, the effect we found of family history of overweight possibly represent a genetic factor or an environmental exposure related to the family.

Additional studies on the relative effect of maternal pre-pregnancy weight and infant birth weight on childhood and especially adulthood weight are needed. Furthermore, studies into the effect of interventions during pregnancy to prevent high birth weight are of interest.

## CONCLUSION

Childhood obesity is a multifactorial disease with complex etiology. Maternal weight is an important, possibly modifiable risk factor. In this study, the strongest risk factors for development of childhood overweight and obesity was maternal obesity and macrosomia (birth weight > 4500g). Social factors such as parents not living together and a family history of overweight or obesity are important, though more complex, predictors. Smoking and breast feeding do not have the same impact in our study as previously reported elsewhere. Further understanding of the underlying biological mechanism for childhood overweight and obesity is needed to be able to design future interventions and give clinical recommendations on how to prevent childhood obesity.

## Data Availability

Some of the data - e.g. the questionnaires - are available upon reasonable request to the authors, whereas other data are restricted.

## ABBREVIATIONS

AGA: Appropriate for gestational age
aOR: Adjusted odds ratio
BMI: Body mass index
CI: Confidence interval
GDM: Gestational diabetes mellitus
IQR: interquartile range
LGA: Large for gestational age
SD: Standard deviation
SGA: Small for gestational age

## Acknowledgements

We are grateful for the work of Mary Ngo and Liv la Cour Poulsen who worked with the organization of the questionnaires.

## Author contributions

IT, LK, TL, J-CH and MC initiated the study, IT, MJLH, and EC analyzed the data. IT, EC, LK, and MJLH wrote the first draft of the manuscript, IT, EC, LK, TL, J-CH, MC, PH and CB revised the manuscript critically, and all authors approved the final version of the manuscript.

